# The effectiveness of full and partial travel bans against COVID-19 spread in Australia for travellers from China

**DOI:** 10.1101/2020.03.09.20032045

**Authors:** Valentina Costantino, David J Heslop, C Raina MacIntyre

**Affiliations:** The Biosecurity Program, The Kirby Institute, UNSW Medicine, The University of New South Wales, Australia; The School of Public Health and Community Medicine, UNSW Medicine, The University of New South Wales, Australia

## Abstract

Australia implemented a travel ban on China on February 1^st^ 2020. Partial lifting of the ban is being considered, given the decline in incidence of COVID-19 in China. We modelled three scenarios to test the impact of travel bans on epidemic control in Australia. Scenario one was no ban, scenario two was the current ban followed by a full lifting from the 8^th^ of March 2020, scenario three was a partial lifting of the current ban to allow over 100,000 university students to enter Australia, but not tourists. We used disease incidence data from China and air travel passenger movements between China and Australia, derived from incoming passenger arrival cards. We estimated the true incidence of disease in China using data on expected proportion of under-ascertainment of cases. We used an age specific deterministic model divided in 18 age stratified groups to model the epidemic in each scenario. The modelled epidemic with the full ban fitted the observed incidence of cases well. The modelled epidemic of the current ban predicts 57 cases on March 6^th^ in Australia, compared to 66 observed on this date, however we did not account for imported cases from other countries. The modelled impact without a travel ban implemented on February the 1^st^ shows the epidemic would continue for more than a year resulting in more than 2000 cases and about 400 deaths. The impact of a partial lifting of a ban is minimal, and may be a policy option. Travel restrictions were highly effective for containing the COVID-19 epidemic in Australia and averted a much larger epidemic. The epidemic is still containable if other measures are used in tandem as cases surge in other countries. This research can inform decisions on placing or lifting travel bans as a control measure for the COVID-19 epidemic.

## Introduction

In response to the epidemic of COVID-19, (1) Australia implemented a travel ban from China on February 1^st^ 2020, adding Iran and then South Korea to the ban on February 29^th^ and March 5^th^ respectively. In addition, Australians evacuated from Wuhan and from the Diamond Princess cruise ship were quarantined for two weeks in dedicated quarantine facilities. The ban on travel from China has been periodically reviewed, with lifting of restrictions announced on February 23^rd^ for high school students, who number less than 800. In contrast, over 120000 university students are unable to enter Australia to commence or resume their studies, and a booming tourism industry has ceased.

Travel bans and social distancing measures are effective public health tools to control epidemic diseases (2), and Australia successfully delayed the introduction of the 1918 pandemic by 1 year and reduced the total mortality compared to other countries (3). However, travel bans are not sustainable indefinitely, and a careful risk analysis needs to be done comparing the health and economic consequences of alternative scenarios. The epidemic in China peaked on February 5^th^ and has declined since (4). The risk of importation of COVID-19 cases through travel from an affected country is proportional to the volume of travel from that country and their prevalence of infection at that time point. We aimed to estimate the impact of the implementation of the travel ban on China from February 1^st^ 2020 on the epidemic trajectory in Australia, as well as the impact of lifting the ban completely or partially from the 8 of March.

## Methods

Three scenarios were considered.

1. No travel ban – the epidemic curve if the travel ban was never placed
2. Complete travel ban from February 1st to March 8st, followed by complete lifting ban
3. Complete travel ban from February 1st to March 8st, followed by partial lifting ban (allowing university students, but not tourists, to enter the country)

### Estimation of infected cases coming into Australia from China

The evacuations from Wuhan and the Diamond Princess Cruise ship are not considered in this model, which only examines regular travel between China and Australia. In order to estimate the effectiveness of the travel ban that has been implemented in Australia for travellers from China, we did not consider bans to other countries. We assumed that the chance of cases coming into Australia from China depends from the number of cases in China and the number of travellers to Australia. To estimate the number of people infected that are predicted to enter Australia every two weeks from 20/01 to April, we utilised 2019 air travel passenger movements between China and Australia, derived from incoming passenger arrival cards, with data aggregated monthly and published by the Australia Bureau of Statistics (5). For the purpose of this analysis air movements of passengers between China and Australia were derived from 2019 data. A baseline level of entries into Australia from China was calculated from the total number of entries over the April – Jun 2019 time period and was assumed to represent the baseline arrivals for the purpose of tourism and other business. The seasonal excess of travellers was then calculated by deducting this baseline from the January-March 2019 data. The seasonal excess arrivals were assumed to represent the arrival of international students starting the 2019 study year, which begins in February to March each year. Where travel bans were instituted in this analysis, or lifted, it was assumed that international students unable to enter Australia would return to Australia following the lifting of the ban, 60% in the rest of March and the remaining 40% over the month of April. However, tourists not able to travel during a travel ban were not assumed to enter Australia at a later date. Tourism activity was assumed to recover to baseline levels immediately after the lifting of a travel ban. The daily number of travellers from China to Australia in each month and for each scenario is showed in Table S2 of the supplementary material.

To then calculate the probable number of those that could be infected we used an epidemiological dataset of confirmed cases of COVID-19 in China collected from WHO situation reports (6) and available in our supplementary materials (Table S1). The dataset includes all confirmed cases in China reported from 31/12/2019 to 23/02/2020. We then assumed that notified cases reflect only 10% of the real new infections per day, due to under-reporting, mild cases and asymptomatic infections. This assumption is based on data from Japan (7), which estimated that only 9.2% of cases in China were notified or detected. This estimate is based on testing of all evacuees from Wuhan to Japan and the documented cases in China at the time (7). Furthermore it has been showed that a high proportion of infected people will have very mild symptoms (8) which are unlikely to be reported. We then estimated the possible true epidemic curve. In order to project the future incidence cases in China we used a Poisson regression model to fit data from the 5^th^ (start of the incidence declining) to 23^rd^ of February and estimated the decreasing rate per day (z) as:

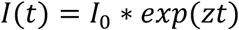

Where I(t) is the number of new infected at time t and I0 is the initial value at time t=0 (Incidence at day 5 of February). Once the decreasing rate z was estimated and the incidence forecasted from 23 of February onwards, we then calculated the number of infected people coming from China every two weeks period *A*(*i, i* + 14) as:

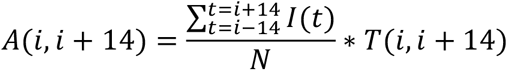

Where N is the total population of China and *T*(*i, i* + 14) is the number of people travelling from China to Australia in every two weeks period. When calculating the prevalence of infection in China, we started from two weeks before the period travelling in order to include the people that could be infected and in a latent state. In scenarios 2 and 3, we assumed a linear declining distribution in time of travel for university students waiting to enter the country after lifting of the travel ban. A full and partial lifting of the ban was examined. In the partial ban, over 150,000 university students can enter Australia, but the just over 80,000 expected tourists not.

### Epidemic curve in Australia from cases imported from China

The cases of COVID-19 occurring over time in Australia due to imported cases from China were estimated for each scenario. We used an age specific deterministic model, with 8 mutually exclusive compartments: susceptible (S), Latent traced (LT), Latent untraced (LU), Infectious (I), Isolated (I), Recovered (R) and dead (D). Each of those compartments is divided in 18 age stratified groups each of 5 years duration, ranging from 0 to 84 years old plus an additional age group of 85+ years. The entire Australian population was considered susceptible. The duration of each model run is 400 days. The initial infected cohort is assumed to be generated from cases arriving from China by air. After arrival of an infected case, it is assumed that, if and when they become symptomatic, they are isolated, and a designated portion of their contacts will also be quarantined. Cases transition between epidemiological compartments in accordance with transition rates determined by their duration of stay in each compartment. Model parameters are shown in Table 1. Further details of the model (diagram and differential equations) are described in the supplementary material. We conducted a sensitivity analyses on the proportion of asymptomatic people, and based on growing evidence of equal viral loads in symptomatic and asymptomatic cases(9-12), we considered the latent period to be equally infectious as the symptomatic period. The proportion of asymptomatic infections, being the main source of community transmission in scenarios where the travel bans are lifted, was assumed to be 34.6% based on testing of passengers aboard the Diamond Princess cruise ship (13, 14). The model uses an optimistic assumption that 80% of contacts are identified and quarantined, and 90% of symptomatic cases are isolated after 5 days (17). Studies show a long, mild prodrome of several days before people feel unwell enough to seek medical attention, which is also considered in the model (15).

**Table 1:**
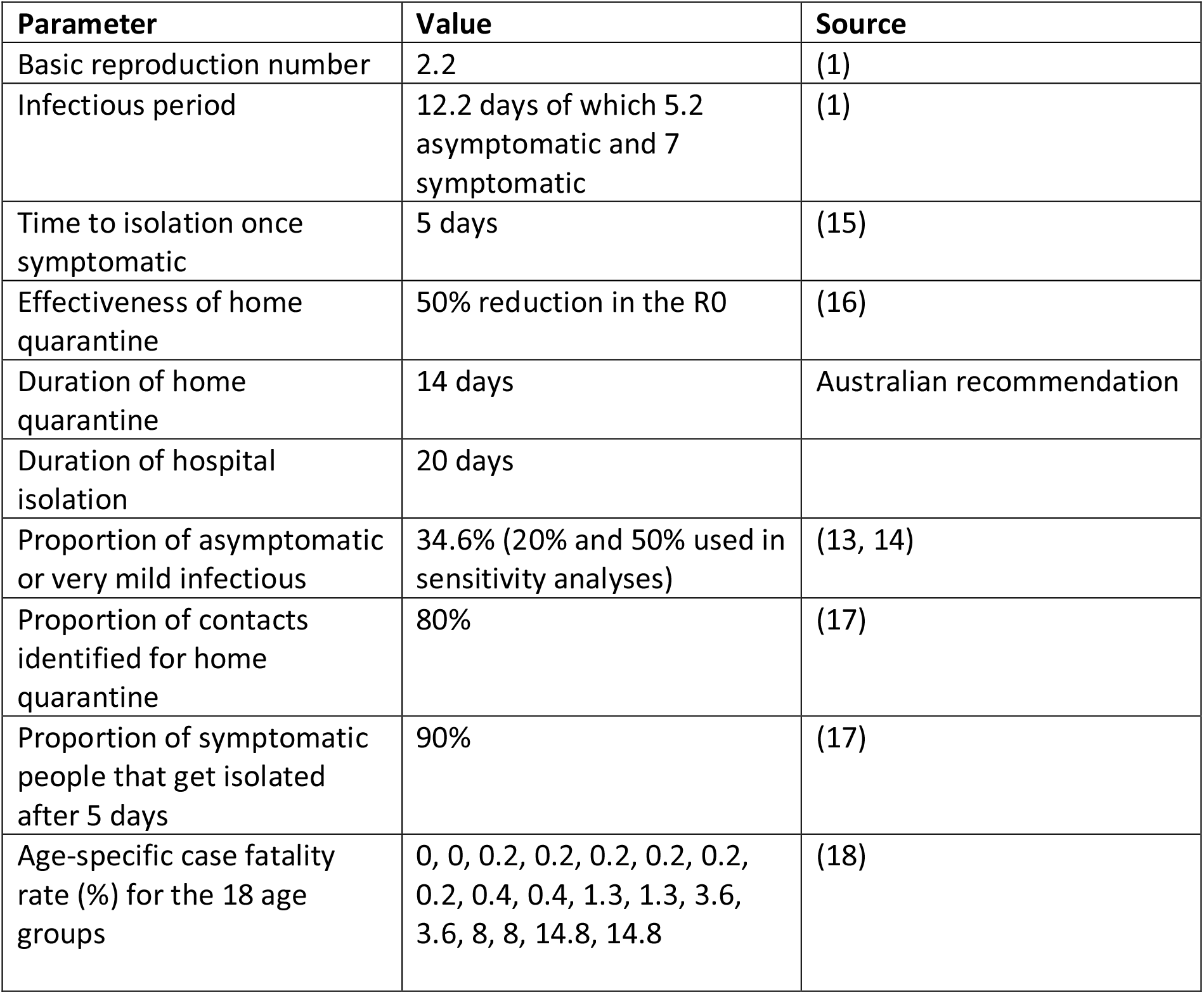
Parameters used in the model

## Results

Figure one shows the notified and estimated epidemic in China from the 31 December 2019 to the 23 of February.

Figure 2 shows the modelled epidemic curve fitting the incidence data from 5 to 23 of February and then forecasted until the 4 of April, which is the time we expect the incidence decreasing to almost zero should the current trend in China continue.

**Figure 1:**
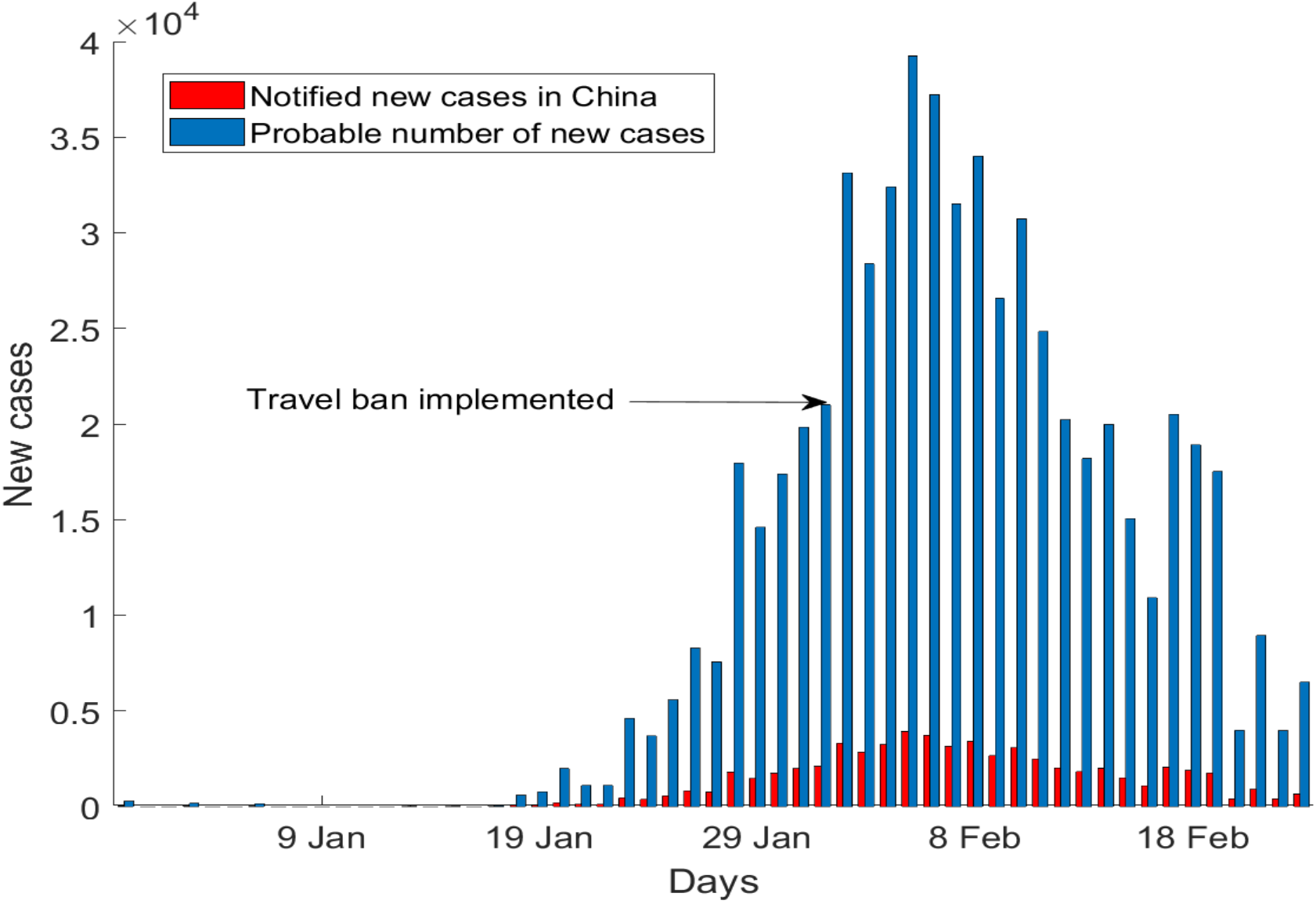
the estimated true epidemic curve (blue) compared to the reported epidemic curve in China (red) (7)

**Figure 2:**
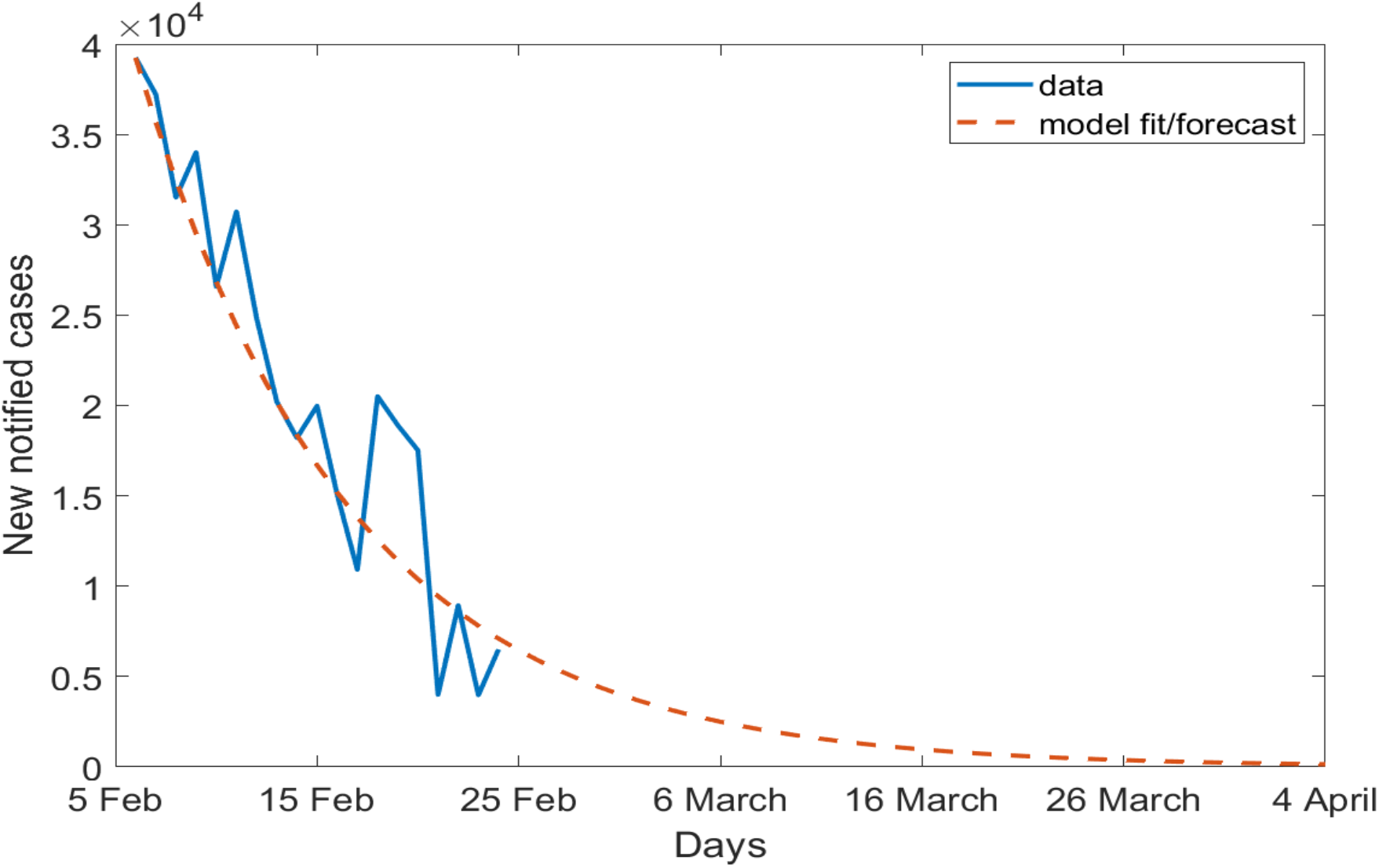
Estimated Incidence data in China (blue) and model fit to the data and forecasting future daily incidence (red)

We found that following the peak on February 5^th^ and decline of the epidemic in China, the probability that an infected traveller can arrive in the partial ban scenario (allowing university students only) is low. The complete removal of travel restrictions on 8^th^ of March results in an estimated arrival of 5 cases in the first two weeks and 1 in the following two weeks. However, if we compare a 5 week ban scenario (scenario 2) with the scenario without a travel ban (scenario 1), we estimate that 32, 43 and 36 infected coming every two weeks from the 26 of January would have been averted. Due to a surge of students coming in the first two weeks following the lifting of the ban in the second scenario, an additional 2 more infected are estimated to enter from 8 to 21 of March (Table 2). In Figure 3 we show the epidemic curve without and with the ban implemented for 5 weeks followed by a full lifting (scenario 1 and 2) and we show a large impact on averting an epidemic in Australia. In both cases, the model reproduces the 15 notified imported cases reported in Australia between the 20 of January and 8 of February. The modelled epidemic in scenario 2, with the full ban, predicts 57 cases in Australia by the 6^th^ of March. The notified cases by 6^th^ of March were 66, however we did not account for imported cases from other countries

**Table 2:** Imported cases in Australia from China under no, partial and full travel bans per each two weeks period.

| **Time travelling** | **Infected entering Australia without ban (scenario 1)** | **Infected entering Australia with ban from 1 February to 7 of March followed by full lifting of ban (scenario 2)** | **Infected entering Australia with following a ban from 1 February to 7 of March followed by partial lifting of ban (scenario 3)** |
| --- | --- | --- | --- |
| 26 January to 8 February | 39 | 7 | 7 |
| 9 to 22 February | 43 | 0 | 0 |
| 23 February to 7 March | 36 | 0 | 0 |
| 8 to 21 March | 3 | 5 | 0 |
| 22 March to 4 April | 1 | 1 | 0 |

**Figure 3:**
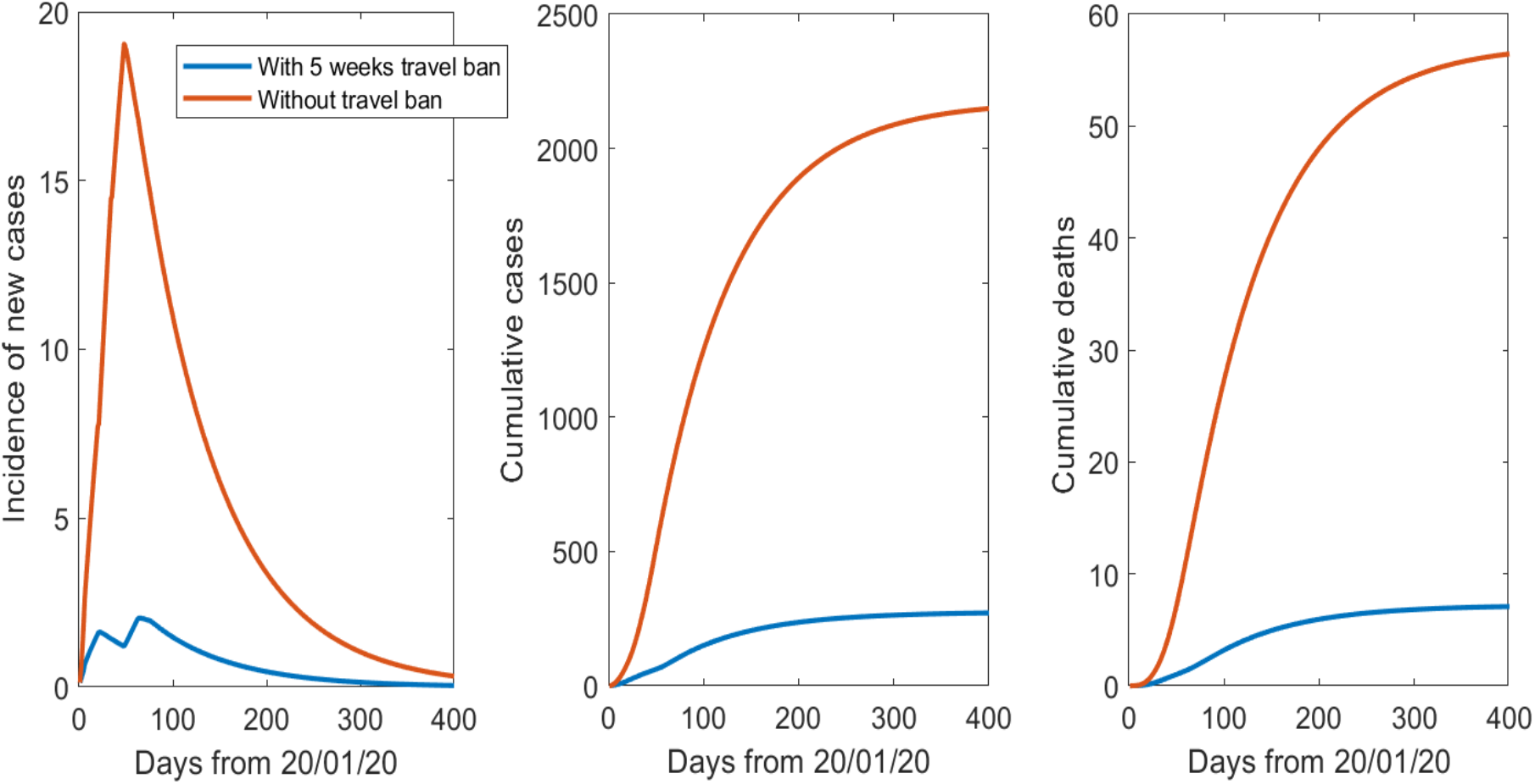
Daily incidence, cumulative number of cases and the cumulative number of deaths from the 20 of January onward for 400 days with and without a travel ban.

In the epidemic curve for scenario 2, when travel resumes there will be a small surge in cases followed by a decrease and the epidemic can be controlled, with a total of less than 300 cases and about 8 deaths. If the ban was never in place (scenario 1), the epidemic would continue for more than a year resulting in more than 2000 cases and about 400 deaths.

Varying the proportion of asymptomatic cases to 20% or 50% keeps about the same percentages of cases and deaths prevented by the ban in the two scenarios, however the total number of cases and deaths are almost 6 times higher and 2 times lower in the case of the proportion of asymptomatic being 50% and 20% respectively (results not shown).

## Discussion

We estimated that the travel ban implemented on 1 of February by Australia has been very effective, reducing the number of cases and deaths from COVID-19 by about 87%. Studies have been published on effectiveness of domestic and international travel restrictions on COVID-19 (19, 20). However, this study is the first one to show the effectiveness of travel ban in Australia, and can inform a phased approach of partial lifting of bans when cases in the source country decline. This allows monitoring of the ongoing situation in China, which may yet see a second wave of the epidemic. Our estimate of the true epidemic curve is supported by other studies (7, 8, 21), and projected case numbers would change with any change in this estimate. Even if the true number of cases in China is 10 or 100 times that reported, only a fraction of the entire population of China has been infected, which leaves a possibility of a subsequent wave of the epidemic. If cases increase in China, the model can provide estimates of risk based on daily new case numbers.

We do not consider cases coming in from other countries – however, this study illustrates the principle of travel bans and public health impact on epidemic control using China as a case study. A further limitation is the uncertainty of parameters used, particularly the proportion of asymptomatic cases. We have used a conservative estimate, but if the rate is higher than 40%, the outcomes would be worse. While it has been showed that distancing measures are highly effective (2, 22) a systematic review looking at the effectiveness of travel restrictions (23), shows that international travel restriction are effective in delaying the epidemic but may not contain it. We also assumed a very optimistic scenario of 80% of contacts being identified, which may not occur with high case numbers, if a high proportion of asymptomatic transmission is occurring, or if self-quarantine is ineffective. In this study we assumed voluntary home quarantine, which is showed to be about 50% effective in R0 reduction (16), however there could be an increased risk of intra-household transmission infected people to contacts (24), which is not considered in this model.

We showed that the ban implemented for travellers from China, when the epidemic was almost at its peak, substantially delayed the spread into Australia. There is now evidence of community transmission in Australia, but the epidemic is still in the early stages, and this study provide evidence to support the new travel bans that have been implemented on Iran and South Korea, in order to delay the epidemic. The model predicted 57 cases by March 6^th^ in Australia, which is slightly less than the notified number of 66, which suggests the model assumptions were reasonable, given we did not account for cases coming in from other countries. Community transmission in Australia in early March is likely linked to imported cases from China, given the fairly long incubation period and less than 3 incubation periods since the first evacuation of Australians from Wuhan on February 3rd. The model fit to observed data was good, also suggesting the epidemic is still possible to contain, if adequate resources are available for thorough contact tracing.

This analyses is a first insight into the effectiveness of travel restrictions for COVID-19 outbreak, supports the effectiveness of the Australian response, informs gradual lifting of the bans or placing of new bans on other countries, and could inform other countries in reducing the burden of importations and resulting domestic transmission of COVID-19.

## Data Availability

Data are available and will be added as supplementary material in a peer-reviewed publication.

https://www.who.int/docs/default-source/coronaviruse/situation-reports/

## Contributions

Valentina Costantino: Methodology and modelling construction, parameterisation of model, writing and revision

David Heslop: Manipulation of travelling data, writing and revision

Raina MacIntyre: Conception of the study and scenarios, parametrisation of model, writing and revision.

All: Designing and conceptualising the model

